# Investigating the Effect of Cardiovascular Exercise on Brain-Derived Neurotrophic Factor in Early Subacute Stroke

**DOI:** 10.1101/2025.03.27.25324771

**Authors:** Bernat De Las Heras, Lynden Rodrigues, Jacopo Cristini, Eric Yu, Ziv Gan-Or, Nathalie Arbour, Alexander Thiel, Ada Tang, Joyce Fung, Janice J Eng, Marc Roig

## Abstract

**Background.:** Following stroke, a growth-promoting response resulting in heightened neuroplasticity occurs during the early subacute stages of recovery, a period during which the brain may be more responsive to therapeutical interventions. Given its central role in regulating neuroplastic processes and brain repair in animal models, brain-derived neurotrophic factor (BDNF) has been investigated as a potential biomarker for stroke recovery in humans, with interventions upregulating it holding therapeutical potential. Cardiovascular exercise (CE) has been recommended for stroke rehabilitation, partly due to its potential to induce neural adaptations, including upregulation of BDNF.

**Objectives.:** To examine the effects of CE on BDNF in individuals at early subacute stages of recovery.

**Methods.:** 76 participants within 3 months of first-ever ischemic stroke were randomly assigned to eight weeks of either CE plus standard care or standard care alone. To measure the chronic and acute responses to exercise in serum BDNF levels, blood samples were collected before and immediately after a graded exercise test conducted at baseline, four and eight weeks. The potential role of the BDNF Val66Met polymorphism in modulating the BDNF response to CE was also explored.

**Results.:** Despite significant increases in cardiorespiratory fitness, CE did not induce any significant chronic or acute changes in BDNF concentration. Similarly, the BDNF response to CE was not modulated by the Val66Met polymorphism or associated with changes in cardiorespiratory fitness and clinical outcomes.

**Conclusions.:** These findings indicate limited effects of CE in modulating circulating BDNF in subacute stages of stroke recovery.

**Trial Registration:** Exercise and Genotype in Sub-acute Stroke: https://clinicaltrials.gov/study/NCT05076747

## INTRODUCTION

Following stroke, there is a time-limited window of heightened neuroplasticity and responsiveness to training during the initial weeks of recovery.^1^ In animal models, this critical period extends for one month, during which a growth-promoting phase propitiates profound functional and structural changes in the brain that underlie both spontaneous and treatment-induced recovery.^2^ These changes include processes such as changes in gene expression, neural excitability, dendritic spine turnover, axonal sprouting, and remapping of neural networks, all regulated by growth-promoting molecules.^2^

In humans, this critical period of neural malleability, known as the early subacute period of stroke recovery, is estimated to occur within the first week to 3 months post-stroke.^3^ During this early stage of recovery, wherein nearly all restoration from impairment occurs, motor rehabilitation interventions appear to induce significantly greater recovery gains compared to therapeutical interventions initiated in later stages.^4^ Nevertheless, unlike in animal models, the neurobiological mechanisms underlying this critical period of recovery in individuals post-stroke remain poorly understood.

Brain-derived neurotrophic factor (BDNF), the most abundant neurotrophin in the brain, plays a crucial role in neural repair by binding to the TrkB receptor and initiating intracellular signaling that drives functional and structural neural changes.^5^ Rodent studies of post-ischemic lesions have demonstrated BDNF’s protective and restorative actions, including mitigating cell death, facilitating synaptic plasticity, and improving functional recovery.^6^ Specifically, BDNF has been shown to mediate early post-stroke motor recovery by regulating synaptic plasticity, underscoring its potential as a biomarker for stroke recovery.^6, 7^

Genetic factors play a significant role in stroke recovery by influencing neuroplasticity and neural repair mechanisms.^8^ The BDNF Val66Met polymorphism, a common variant of the BDNF gene involving a substitution of valine (Val) for methionine (Met) at codon 66, have been associated with reduced post-stroke recovery, likely due to its potential to interfere with activity-dependent BDNF secretion.^9^ Studying the BDNF Val66Met polymorphism and its impact on BDNF protein levels could enhance our understanding of the neurobiological processes underlying stroke recovery.

BDNF is an activity-dependent neurotrophic factor, with its expression, secretion, and action susceptible to interventions with the potential to enhance neural activity.^10^ Cardiovascular exercise (CE) is a simple yet effective intervention to protect and maintain brain function through its capacity to promote key neurobiological processes within the nervous system.^11^ In animal models, CE interventions have been shown to support functional recovery after stroke, in part by stimulating processes such as synaptic plasticity and upregulation of neurotrophic factors, including BDNF, with BDNF playing a crucial role in mediating CE-induced recovery after stroke.^12^

In neurotypical individuals, CE transiently increases circulating BDNF concentrations following a single bout of CE (BDNF_acute_) and, less consistently, after chronic interventions involving multiple bouts of CE (BDNF_chronic_).^13^ Additionally, studies in both animals and humans have suggested amplified BDNF_acute_ responses following chronic CE programs, indicating an increased BDNF_acute_ responsiveness induced by CE training.^14^ Reduced BDNF levels, which have been observed in patients with stroke, have been associated with poorer long-term functional outcomes.^15^ Similar to neurotypical individuals, though with more inconsistent findings, CE interventions after stroke have shown the potential to modulate circulating BDNF levels, with increases reported both following a single session and training programs.^16^

All existing studies, however, focused on patients within chronic stages of stroke recovery (>6 months),^17^ thereby neglecting the critical period during which the brain might be more responsive to training.^1^ To address this gap, we conducted a study to evaluate the effects of CE on circulating BDNF levels in individuals <3 months of post-stroke recovery. Over an 8-week period, during which participants received either CE+standard care or standard care alone, BDNF_chronic_ levels were measured at rest and BDNF_acute_ after a single CE session. We also examined the associations between BDNF responses and changes in recovery outcomes, as well as the potential influence of Val66Met. We hypothesized that CE would increase both BDNF_chronic_ and BDNF_acute_ response and that carrying the Val66Met would attenuate the response.

## METHODS AND MATERIALS

### Design

In this registered randomized controlled trial (NCT05076747) participants were assigned to either an 8-week CE training in addition to standard care or standard care alone (**Figure 1**). Given the unequal allelic frequency of the Val66Met in different populations,^18^ the randomization sequence allocated more participants in the CE training group to increase the power for detecting effects of this polymorphism on the BDNF response to CE. Assessments occurred at baseline (T0), four weeks (T1), and eight weeks (T2). Each assessment consisted of two experimental sessions 48 hours apart, comprising clinical motor outcomes and cardiorespiratory fitness with blood collection for BDNF. Information regarding participant’s characteristics and relevant clinical information were collected at T0. Enrollment occurred between June 2018 and July 2023. The site ethics board approved the study (Centre de Recherche de Readaptation du Montréal, CRIR-1265-0817) and all participants provided written informed consent.

**Figure 1.**
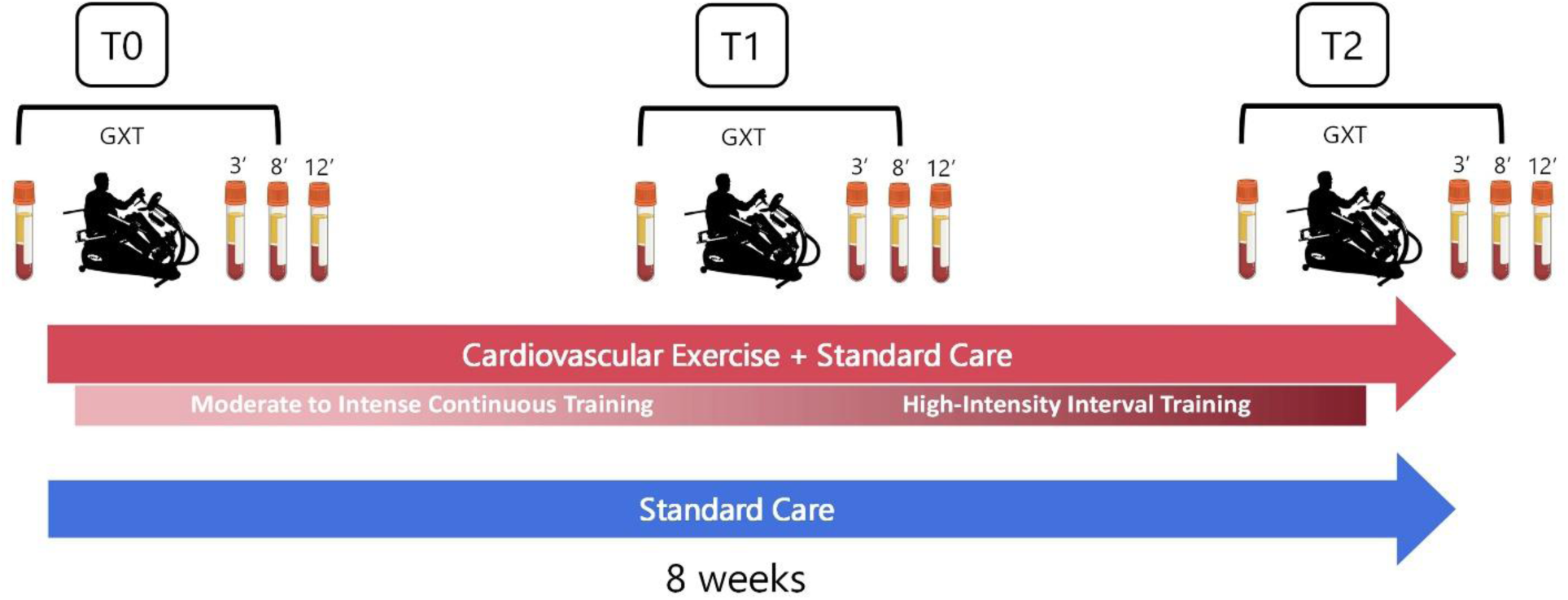
Study design with blood collection evaluations at baseline (T0), four weeks (T1), and eight weeks (T2). Participants were randomly assigned to either an 8-week CE training in addition to standard care (red arrow) or standard care alone (blue arrow). The 8-week CE intervention consisted of 4 weeks of moderate to intense continuous training followed by 4 weeks of high-intensity interval CE training. To measure the effects of CE training on circulating BDNF_chronic_, blood samples were taken at rest before the graded exercise test (GXT). To measure BDNF_acute_, blood samples were taken at 3, 8 and 8 minutes after the GXT. These measurements were conducted in both groups at each time point (T0-T2). Abbreviations: CE, cardiovascular exercise; GXT, graded exercise test.

### Participants

We only included participants with first-ever ischemic stroke within the early subacute stages of recovery (7 days-3 months). Participants had to be between 40 and 80 years old, presented with no upper-limb musculoskeletal or neurological conditions other than stroke, had sufficient ability/capacity to perform the CE training and assessments safely, and had sufficient cognitive/communicative capacity to understand instructions. Individuals were excluded if they had a hemorrhagic stroke, cognitive impairment/dysphasia affecting informed consent, absolute contraindications to exercise,^19^ or were concurrently enrolled in another CE training program.

### Assessments

#### Baseline Assessments

At baseline (T0), stroke severity and cognitive status were assessed with the National Institutes of Health Stroke Scale (NIHSS) and the Montreal Cognitive Assessment (MoCA), respectively. The age-adjusted Charlson Comorbidity Index (CCI) was employed to assess pre-existing comorbidities. Self-reported physical activity levels were measured at each time point using the physical activity scale for people with disabilities (PASIPD). Participants were instructed not to engage in moderate- or high-intensity physical activity 24 hours before the assessments.

#### Cardiorespiratory Fitness

Measurement of peak oxygen uptake (VO_2_peak in mL.Kg^-1^.min^-1^) with a graded exercise test (GXT) is the gold standard for determining cardiorespiratory fitness. A symptom-limited GXT utilizing a protocol validated for individuals with stroke was performed on a whole-body recumbent stepper (NuStep T4r, Michigan, USA).^20^ During the GXT, heart rate (HR) was measured continuously while blood pressure (BP) and rate of perceived exertion (RPE) were recorded every 2 minutes. The GXT was also used to determine maximal HR (HR_max,_ beats per minute -bpm-) and peak power output (PPO, Watts -W-). Indications for test termination followed current guidelines.^21^

#### Clinical Motor Outcomes

Upper-limb motor impairment was evaluated using the Upper-Limb Fugl-Meyer Assessment (UL-FMA), while changes in upper-limb function on the affected side were assessed with the Box and Block Test (BBT).

#### Blood Collection and Analysis

Blood collection was carried out by a nurse, with patients instructed to abstain from eating for at least two hours before the GXT. An antecubital intravenous line was placed in the non-paretic arm, with a waste sample collected before each blood extraction and the line flushed after each draw. A 5 mL blood sample was collected in a vacutainer serum separator tube 10 minutes before the GXT and at 3, 8, and 12 minutes post-GXT to assess BDNF_chronic_ and BDNF_acute_, respectively.^22^ BDNF_acute_ was calculated as the change between pre-GXT BDNF level, and the average concentrations measured at 3, 8, and 12 minutes post-GXT (**Figure 1**).

It was not possible to perform blood collection at the same time for all participants, but for each participant, samples were collected at the same time at T0, T1 and T2. Upon collection, blood samples were clotted for 1 hour, resting at room temperature, followed by 30 minutes at ∼4⁰C, and then centrifuged at 2200g for 15 minutes. The resulting serum was then aliquoted into 250μL cryovials and stored in a -80⁰C freezer. Identified as one of the best-performing assays,^23^ the Biosensis Mature BDNF Rapid^TM^ enzyme-linked immunosorbent assay (ELISA) Kit was employed to determine BDNF concentrations.

#### Genotyping

Genomic DNA was extracted from red blood cells or saliva samples (DNA Genotek Inc., Canada) collected at baseline (T0), and genotyped using the Infinium Global Diversity Array-8 v1.0 from Illumina. DNA extraction and purification were processed by Genome Quebec (Quebec, Canada) using the QIAsymphony system (QIAGEN). Sixty-eight individuals were genotyped with sufficient DNA concentration for reliable genotyping (10ng/ul). Standard quality control was performed using PLINK v1.9. Subjects were classified based on their genotype for the BDNF single nucleotide polymorphism rs6265 as homozygous for the Val allele (Val/Val), heterozygous (Val/Met), and homozygous for the Met allele (Met/Met) using PLINK v1.9 **(Table 1)**. Individuals with Val/Met and Met/Met genotypes were combined to increase statistical power.^24^

**Table 1.** Baseline demographic and clinical outcomes. Values are presented as mean ± SD. Abbreviations: AC, anticoagulant; ACE, Angiotensin-Converting Enzyme; AP, antiplatelet; BB, beta-blocker; BBT, Box and Block Test; BMI, body mass index; CCI, Charlson Comorbidity Index; CE, cardiovascular exercise; F, female; M, male; Met, methionine; METs, metabolic equivalent of task; MoCA, Montreal Cognitive Assessment; NIHSS, National Institutes of Health Stroke Scale; PSY, psychoactive; SNP, single-nucleotide polymorphism; STA, statin; UL-FMA, upper-limb Fugl-Meyer; Val, Valine.

|  | **CE+Standard Care**<br>**(n= 48)** | **Standard Care**<br>**(n= 28)** |
| --- | --- | --- |
| **Age (years)** | 63 ± 11.39 | 65.35 ± 8.68 |
| **Sex (F/M)** | 11/37 | 10/18 |
| **BMI** | 27 ± 3.46 | 26.52 ± 4.01 |
| **Time since stroke (days)** | 68.12 ± 22.07 | 58.75 ± 24.03 |
| **Lesion location (%)** |  |  |
| **Cortical** | 16 | 21 |
| **Cortico-subcortical** | 19 | 14 |
| **Subcortical** | 54 | 57 |
| **Cerebellar/Brainstem** | 10 | 7 |
| **NIHSS (0-42)** | 2.02 ± 2.20 | 1.92 ± 1.92 |
| **MoCA (0-30)** | 24.14 ± 4.85 | 23.21 ± 3.8 |
| **UL-FMA (0-66)** | 56.20 ± 10.24 | 59.14 ± 8.22 |
| **BBT<sub>affected</sub> (blocks/min)** | 46.8 ± 13.25 | 48.10 ± 12.65 |
| **Cardiorespiratory fitness**<br>**(VO<sub>2</sub>peak, mL.Kg<sup>-1</sup>.min<sup>-1</sup>)** | 17.88 ± 5.49 | 18.12 ± 5.58 |
| **SNP rs6265** |  |  |
| **Val/Val** | 27 | 18 |
| **Val/Met** | 13 | 7 |
| **Met/Met** | 3 | 0 |
| **CCI (Age-adjusted)** | 4.57 ± 1.83 | 4.57 ± 1.66 |
| **Walking aid dependence (%)** | 15 | 13 |
| **Smoking history (%)** |  |  |
| **Non-smoker** | 52 | 43 |
| **Former smoker** | 40 | 56 |
| **Current smoker** | 8 | 1 |
| **Medications (n)** | 5.07 ± 2.58 | 5 ± 2.22 |
| **Classification (%)** |  |  |
| **AC** | 60 | 53 |
| **ACE** | 31 | 42 |
| **AP** | 46 | 60 |
| **BB** | 35 | 25 |
| **PSY** | 33 | 28 |
| **STA** | 79 | 100 |
| **Therapy sessions (n)** |  |  |
| **Physiotherapy** | 8.87 ± 8.21 | 6.59 ± 5.78 |
| **Occupational Therapy** | 11.5 ± 8.26 | 7.45 ± 5.98 |
| **Speech Therapy** | 5.02 ± 8.81 | 2.15 ± 5.48 |
| **ΔT0-T2 Physical activity (METs**<br>**hr/day)** | 1.48 ± 5.39 | -0.23 ± 5.28 |

### Intervention

#### Cardiovascular Exercise

The CE+standard care group underwent a total of 24 CE training sessions over an 8-week period, with a frequency of 3 times a week and a 48-hour rest between sessions whenever possible. CE comprised four weeks of progressive moderate-to-vigorous intensity continuous training (MICT) followed by four weeks of progressive high-intensity interval training (HIIT), all conducted on a whole-body recumbent stepper ergometer **(Figure 1)**. Each training session included 2.5 minutes of warm-up and cool-down at 35% of the PPO, along with the main training component at the targeted intensity. Blood pressure was measured at the beginning and the end of each CE session. To quantify the CE stimulus, HR, and Watts were continuously monitored during training via a pulse sensor (Polar H10, Kempele, Finland) and the stepper’s digital console, respectively. RPE (0-10) was assessed every 5 minutes throughout each training session with the modified Borg scale. Training variables, including the average percentage of maximal HR (%HR_max_), the average percentage of PPO (%PPO), total steps, and average RPE, were calculated for each session to quantify internal and external training workloads.

##### Moderate-to-vigorous Continuous Training (weeks 1-4)

MICT has been typically employed as a standard CE modality in stroke rehabilitation programs. Intensities were determined using the PPO associated with VO_2_peak during the GXT at T0 and progressively increased by 5% weekly from 65% to 80% PPO, to promote training adaptations. Session durations also increased from 20 to 35 minutes.

##### High-intensity Interval Training (weeks 5-8)

HIIT intensities were determined using the PPO corresponding to the VO_2_peak level achieved during the GXT at T1. The HIIT protocol comprised 8 x 60-second high-intensity intervals (8 minutes) interspersed with 7 x 60-second low-intensity intervals (7 minutes), totaling 20 minutes per session. This 60:60 interval ratio is optimal for sustaining high intensities.^25^ While high-intensity intervals began at 85% PPO and increased by 5% weekly until reaching 100% PPO, low-intensity intervals were kept constant at 35% PPO. To minimize sudden changes in BP while ensuring target intensities, the workload was progressively increased (15 seconds) before each high-intensity interval.

### Standard Care Program

Standard care consisted of rehabilitation sessions conducted in the same center as the intervention and prescribed by the stroke clinical unit. In addition to routine health monitoring by physicians and nursing staff, standard care included physiotherapy, occupational therapy, and speech therapy sessions. The content, duration, and amount of rehabilitation varied among patients and were tailored to individual needs as determined by the stroke clinical unit, with each therapy session lasting 45 minutes. To examine potential differences between groups in standard care, we recorded the type and number of therapy sessions received by each patient from the beginning of the study to its conclusion.

### Statistical Analysis

Data were plotted using normality plots and histograms for inspection. The Shapiro-Wilk test was used to confirm normality for each variable. Baseline differences in participant characteristics and clinical variables between groups were assessed using t-tests or Wilcoxon tests. Linear mixed models (LMM) were used to analyze differences in clinical motor outcomes (UL-FMA, BBT), cardiorespiratory fitness, and BDNF measures between groups across time points (T0-T2). Each model included either BDNF_chronic_ or BDNF_acute_ as the dependent variable, with time point (T0-T2), group, and their interaction as fixed effects. Covariates in the model included age, sex, and stroke severity (NIHSS). Body mass index was also entered into the model as a covariate due to its significant effect on BDNF levels.^26^ Participants were treated as a random effect to account for individual differences at baseline. Exploratory analyses combining data from both groups were conducted to measure BDNF_acute_ at baseline (T0). The Tukey’s HSD test was applied to identify statistically significant pairwise differences.

To examine the potential influence of Val66Met (Val/Val vs. Val/Met + Met/Met), the allele variant was nested within the Time x Group interaction. Based on the Bayesian Information Criterion, log-likelihood ratio tests, and the data’s temporal dependence, an AutoRegressive (AR1) covariance structure was deemed most appropriate. Assumptions for linear models, including normality in the distribution of random coefficients, were examined for all the variables in the model.

Standard least squares multivariate linear regression analyses were used to investigate associations between BDNF_chronic_ and BDNF_acute_ with changes in cardiorespiratory fitness and clinical motor outcomes. The same covariates—age, sex, stroke severity, and BMI-were included. Multicollinearity between predictor variables was assessed with the variance inflation factor (VIF) with a threshold of ≤5, indicating unacceptable multicollinearity.^27^ All statistical analyses were performed with JMP (SAS Institute Inc, Cary, NC), version 17, and tested for significance at 0.05 alpha level (p<0.05).

## RESULTS

**Table 1** presents the participant’s characteristics and relevant clinical information for both groups at baseline. The trial flow, including dropouts, is detailed in **Figure 2**. Data from all participants were included to measure BDNF_acute_ responses at T0, and an intention-to-treat approach was used for those who were assessed at least at T1. No adverse events related to training were reported.

**Figure 2.**
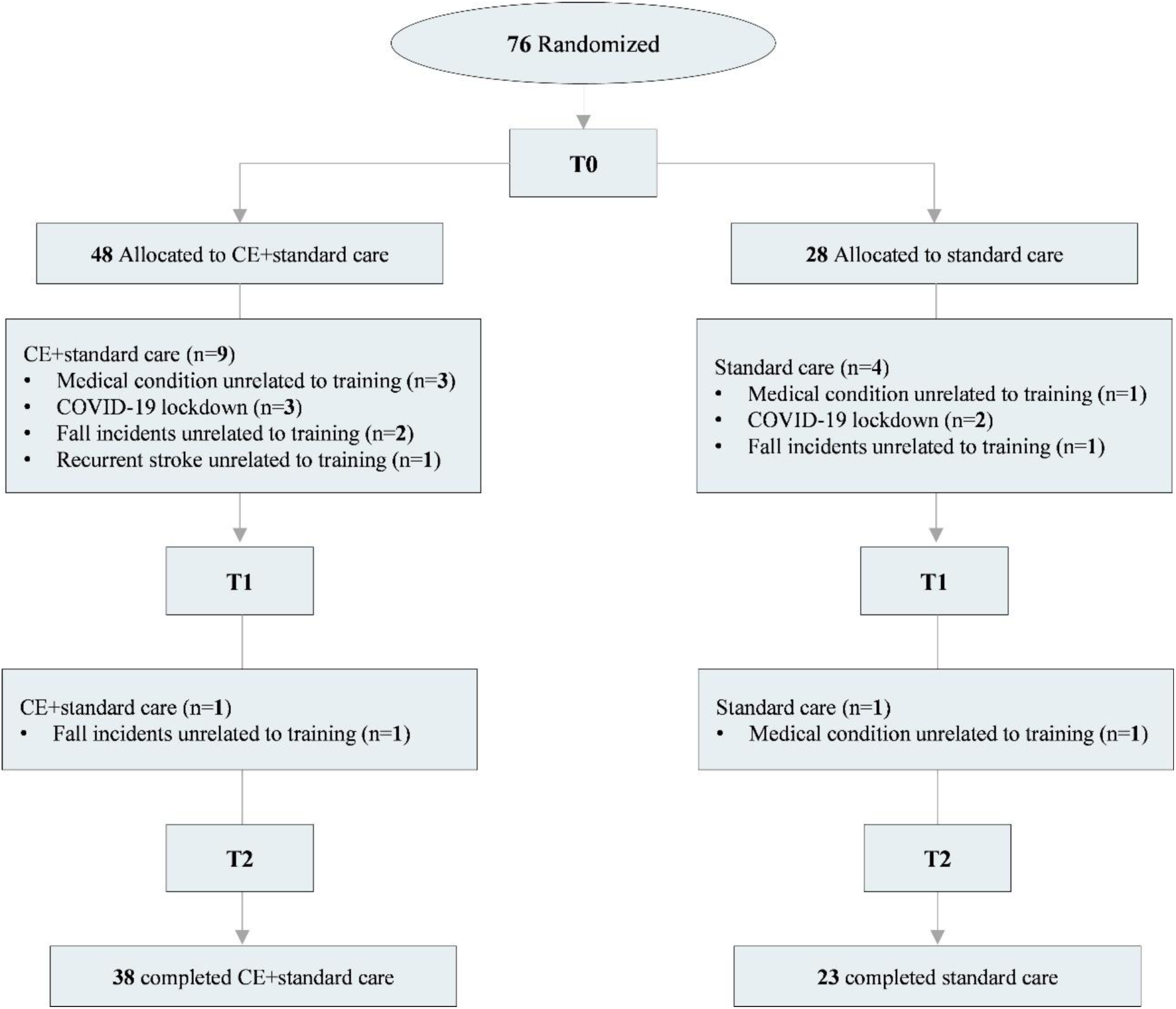
Flow chart of the Randomized Controlled Trial. Abbreviations: CE: cardiovascular exercise; n: number of participants; T0: baseline; T1: four weeks; T2: eight weeks.

On average, participants were 63.5±10.2 years old (mean ± SD) and initiated the study 65.1±22.8 days after stroke. Participants presented mild stroke severity, with an average NIHSS score of 2.01±2.09, and an average MoCA score of 23.8±4.48. No significant differences were observed at T0 between groups in terms of age, sex, body mass index (BMI), time since stroke, lesion location, stroke severity, cognitive status, upper-limb impairment and function, pre-existing comorbidities (measured with age-adjusted CCI), walking aid dependence, smoking history, and the average number of prescribed medications. The amount of standard care provided during the participation in the trial and levels of physical activity outside of the trial were similar between groups. All participants assigned to the CE+standard care group who completed the study attended all 24 sessions. Internal and external training workloads during CE training are reported in **Supplementary Table 1.**

### Cardiorespiratory Fitness

No significant differences in cardiorespiratory fitness were observed between groups at baseline (T0). At T0, all participants had an average VO_2_peak of 18.43±5.63, a HR_max_ of 81±13% of the age-predicted maximum, and an average time to exhaustion of 10.49±2.50 minutes **(Table 2)**. There was a significant effect of Time (F(2,78) = 16.76, p = <.0001), and a significant Time x Group interaction (F(2,78) = 13.46, p = <.0001). While the standard care group showed no significant change in VO_2_peak from T0 to T2 (+0.27 mL.Kg^-1^.min^-1^, 95% CI -2.19 to 1.64, p=0.998). The CE+standard care group showed significant VO_2_peak improvements, with an increase of +2.76 mL·kg⁻¹·min⁻¹ (95% CI: 1.58 to 3.93, p < .0001) at T1 during MICT, further increasing by +1.64 mL·kg⁻¹·min⁻¹ (95% CI: 0.45 to 2.82, p < .0001) at T2 following HIIT, for a total gain of 4.43 mL·kg⁻¹·min⁻¹ from T0 (95% CI: 2.97 to 5.82, p < .0001).

**Table 2.**
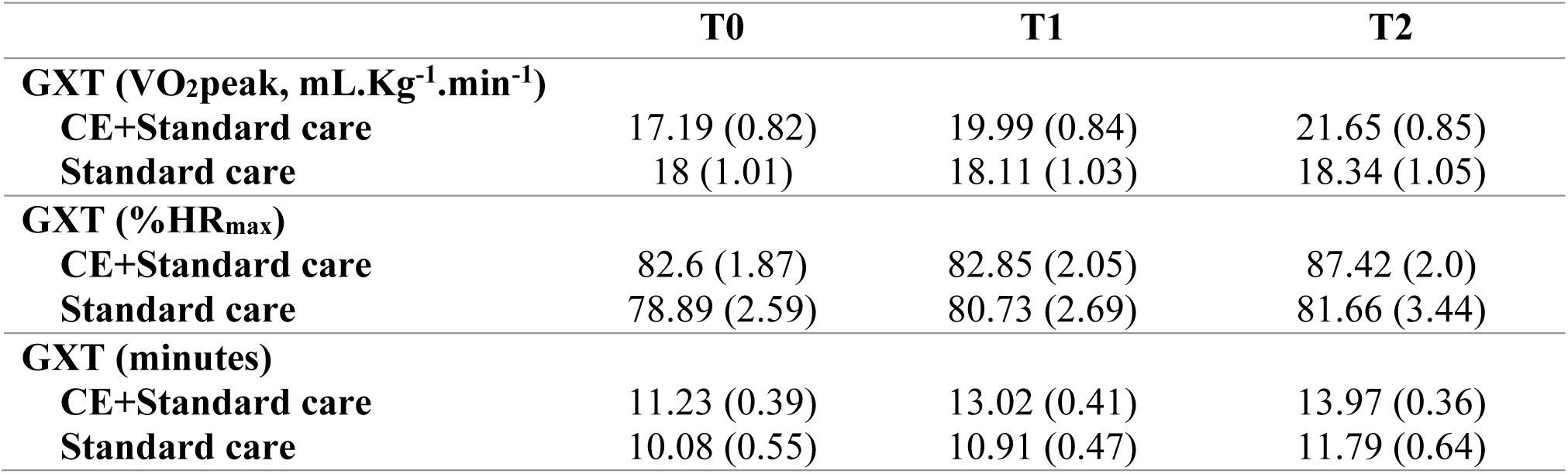
GXT values at baseline (T0), four weeks (T1), and eight weeks (T2) for the CE+standard care and standard care groups. Measurements include maximum oxygen uptake (VO_2_peak), %HR_max_, calculated based on the HR relative to the age-predicted maximum. and average time to exhaustion in minutes. Data are presented as least squares means with standard errors (SE). Abbreviations: CE, cardiovascular exercise; GXT, graded exercise test; HR, heart rate; mL.Kg^-1^.min^-1^, milliliters per kilogram per minute.

### Clinical Motor Outcomes

There was a significant effect of Time on upper-limb motor impairment and function using the UL-FMA (F(2, 99)=15.61, p=<.0001) and BBT (F(2, 116)=15.73, p=<.0001), with no significant Time x Group interaction for either measure (UL-FMA: F(2, 99)=1.04, p=0.355; BBT: F(2, 116) =0.22, p=0.801).

### BDNF Concentration

Two participants did not go through blood sample collection, resulting in no BDNF data being available for analysis. BDNF_chronic_ and BDNF_acute_ changes for both CE+standard care and standard care groups are detailed in **Table 3**. At T0, no statistically significant differences in basal BDNF concentration were observed between groups (p= 0.275). Similarly, no significant effects of Time (F(2,186) = 1.08, p = 0.340) or Time x Group (F(2,186) = 0.06, p = 0.937) were identified for BDNF_chronic_ **(Figure 3A)**.

**Table 3.** BDNF_chronic_ and BDNF_acute_ concentration at baseline (T0), four weeks (T1), and eight weeks (T2) following CE+standard care and standard care groups. BDNF_chronic_ was assessed by comparing the basal concentrations at rest across the study time points, while BDNF_acute_ was determined as the difference between resting levels pre-GXT and the average concentration levels post-GXT (3, 8, and 12 minutes). Data are presented as mean and SD. Abbreviations: CE, cardiovascular exercise; GXT, graded exercise test; pg/mL: picograms per milliliter.

| Serum concentration (pg/ml) | **T0** | **T1** | **T2** |
| --- | --- | --- | --- |
| **BDNF<sub>chronic</sub>** |  |  |  |
| **CE+Standard care** | 23260 ± 6433 | 24084 ± 7554 | 24809 ± 6774 |
| **Standard care** | 25094 ± 7682 | 25946 ± 8586 | 26536 ± 8983 |
| **BDNF<sub>acute</sub>** |  |  |  |
| **Δ CE+Standard care** | 233 ± 4060 | -260 ± 3037 | 156 ± 3144 |
| **Δ Standard care** | -507 ± 2821 | -2173 ± 3622 | -100 ± 3414 |

**Figure 3.**
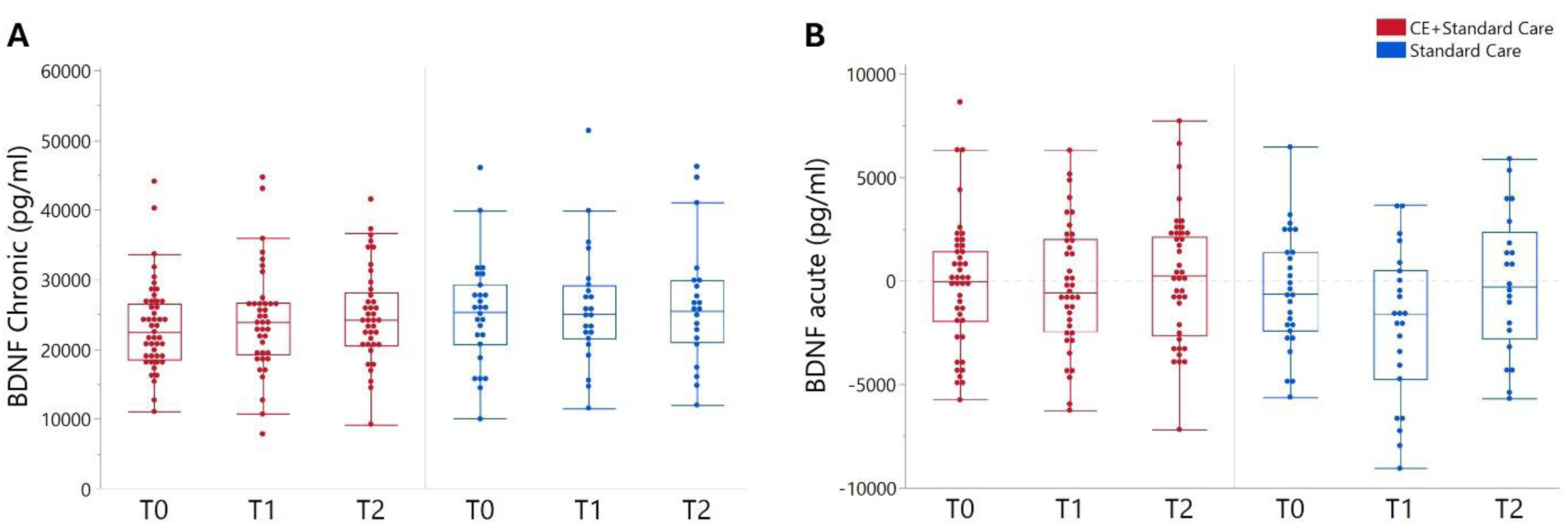
BDNF_chronic_ (A) and BDNF_acute_ (B) changes in BDNF concentration at baseline (T0), four weeks (T1), and eight weeks (T2) following CE+standard care and standard care groups. Data are presented as raw values, with whisker plots representing interquartile range and potential outliers. Abbreviations: CE, cardiovascular exercise, pg/mL: picograms per milliliter.

No significant effects of Time (F(2,184) = 2.76, p= 0.065) or Time x Group (F(2,184) = 1.01, p= 0.364) were identified on BDNF_acute_ throughout the study (T0-T2) (**Figure 3B**). In the exploratory analysis combining both groups at T0 (n=74) significant Time effects on BDNF_acute_ were observed (F(3,282) = 2.67, p = 0.047). Specifically, there was a non-significant increase from baseline to 3 minutes post-GXT (+574.93 pg/ml, 95% CI -672.18 to 1824.03, p=0.632) and significant decrease between 3 and 12 minutes post-GXT (−1345.19 pg/ml, 95% CI -2593 to 96.47, p=0.029).

Our findings showed no significant effects of Val66Met on either BDNF_chronic_ or BDNF_acute_ responses **(Supplementary Table 2)**. Similarly, no significant associations were observed between BDNF_chronic_ and BDNF_acute_ responses and changes in clinical motor outcomes (UL-FMA, BBT) and cardiorespiratory fitness in either the CE+standard care or the standard care group (**Supplementary Table 3**).

## DISCUSSION

Rehabilitative treatments capable of promoting neuroplasticity such as CE are believed to have therapeutic potential for stroke recovery, especially during the early post-injury, stages when the brain may be highly responsive to treatment.^1^ This study is the first to examine the effects of CE on BDNF levels in individuals with early subacute stroke. Contrary to our hypotheses, and despite clinically significant improvements in cardiorespiratory fitness **(Table 2)**, 8-week progressive CE training did not significantly affect BDNF_chronic_ or BDNF_acute_ responses. Furthermore, BDNF responses were neither modulated by Val66Met polymorphism nor associated with clinical motor outcomes following either CE training plus standard care or standard care alone.

CE is a core component of stroke rehabilitation with a well-established capacity to enhance cardiorespiratory health and metabolic function, reducing stroke recurrence risk factors, while also potentially supporting brain function and neural recovery.^12^ Studies on rodents have shown that several days of voluntary CE increase BDNF expression and its receptor TrkB in the brain, a molecular response mediating activity-dependent neuroplasticity processes supporting learning and memory, as well as neural repair post-stroke.^28^ However, the effects of CE on neuroplasticity and brain repair in individuals after stroke remain largely unknown, especially in the early stages of recovery.^17^

Despite the effectiveness of the CE intervention in improving cardiorespiratory fitness, our findings revealed no significant effects on BDNF_chronic_. The CE program led to significant increases in cardiorespiratory fitness, with average VO_2_peak increases of 4.43±3.24 mL.Kg-1.min-1 (+27.25%). These increases exceed the minimal clinical important difference of 3.0 mL.Kg-1.min-1,^29^ and surpass previously reported improvements in both subacute stroke individuals undergoing high-intensity CE interventions (+1.46 mL/kg/min)^30^ and chronic stroke populations in BDNF studies (Ploughman et al., 2019: +1.7 mL/kg/min^31^; Hsu et al., 2021: +3.4 mL/kg/min^32^). It is therefore unlikely that the lack of BDNF_chronic_ increase could be due to an insufficient exercise stimulus. This view was further supported by our regression analysis, which found no significant association between changes in VO_2_peak and BDNF_chronic_ (**Supplementary Table 3)**.

While unexpected, these results are consistent with the mixed evidence on the impact of CE training on BDNF_chronic_ in humans. In neurotypical populations, studies have presented conflicting findings, with some investigations reporting increased BDNF_chronic_ following long-term CE interventions, while others showing no change or even reductions.^13^ Only two studies have investigated the long-term effects of CE training on BDNF_chronic_ in patients with chronic stroke, with divergent results. One study (n=23) reported significant increases after 12 weeks of HIIT,^32^ whereas another study (n=52) reported no significant changes following 10 weeks of vigorous-intensity treadmill training compared to a group undergoing standard care.^31^

In contrast to other neurotrophins that are secreted constitutively, under resting conditions, BDNF remains within the cytoplasm and is only secreted in response to neural activity.^10^ This activity-dependent release is also evident following CE, where current evidence robustly supports BDNF_acute_ increases following a single CE session but does not consistently show BDNF_chronic_ after long-term training programs.^13^ This distinction could be of importance in stroke recovery, as the transient upregulation of intracellular signaling molecules in the brain like BDNF after a single exercise session initiates a biochemical cascade responsible for synaptic changes previously related to neural repair.^33, 34^ Additionally, although long-term CE may not significantly increase BDNF_chronic_, animal studies suggest that long-term training programs can prime the BDNF_acute_ response to a single exercise session, indicating an adaptive mechanism.^14^

Our findings did not show any significant priming effects of CE training on BDNF_acute_ response, and the exploratory analysis combining both groups demonstrated only moderate effects on BDNF_acute_ in response to a GXT at T0. By using a GXT, we were able to measure BDNF_acute_ responses following a high-intensity CE session while also evaluating its association with cardiorespiratory fitness (VO_2_peak). These findings contrast with previous studies reporting BDNF_acute_ increases following a single vigorous CE session, including a GXT, in both neurotypical populations,^22^ and individuals in the chronic stage of stroke.^35^ Additionally, while studies in neurotypical populations show that several weeks of CE training can enhance BDNF ^36^ our findings align with the only stroke study examining acute responses to training, which found no effect of 10 weeks of vigorous-intensity treadmill training on BDNF_acute_ immediately after a GXT.^31^

One possible contributor to the limited effects of CE on BDNF could be the stress and inflammatory processes that characterize early post-stroke stages. Stroke triggers a cascade of stress-related hormones (e.g. corticosterone, cortisol) and pro-inflammatory molecules (e.g. Interleukin-6, tumor necrosis factor-alpha, or C-reactive Protein), which can persist during the acute and subacute stages,^37, 38^ attenuating BDNF mRNA levels and BDNF expression.^39, 40^ Furthermore, while long-term CE has shown to offer both anti-inflammatory and stress-reducing benefits,^41^ animal studies show that a single CE session, when implemented at higher intensities, can stimulate pro-inflammatory responses and cause up to a 20-fold increase in corticosterone levels, thereby reducing BDNF expression.^33, 42^ Given the high intensities attained during the GXTs in our study (**Table 2**), it is possible that our acute intervention could have acted as a stressor, potentially suppressing any BDNF_acute_ responses in early subacute stages, when growth-inhibiting processes are significant.^1, 43^ These findings are consistent with previous animal work showing that a high-intensity motorized running session implemented two weeks post-stroke resulted in an attenuated BDFN response alongside significantly elevated serum corticosterone levels.^44^

While the inability of CE to promote BDNF, particularly BDNF_acute_ increases, could be related to inhibiting processes characteristic of the early stages of post-stroke recovery, the fact that negative findings have also been reported in chronic stages post-stroke,^31^ suggests that other factors may also contribute. It is important to note that several methodological and biological factors, such as age, BMI, diurnal variations, fasting state, and, importantly, medications like anti-platelets commonly prescribed after stroke, have also been shown to significantly affect circulating BDNF levels.^26, 45^ Although we made efforts to control for these variables, their influence on our findings cannot be entirely ruled out. Addressing these factors, although logistically complex, will be crucial for reducing the significant variability in BDNF measurements and improving both the reliability and replicability of research findings.

In line with previous evidence showing that peripherally measured BDNF has limited predictive recovery post-stroke,^46^ we found no significant associations between BDNF responses and changes in clinical motor outcomes in either CE+standard care or standard care groups **(Supplementary Table 3)**. The lack of associations could be due to many reasons such as the null increase in BDNF in response to CE training, the fact that this intervention had little effect on clinical motor outcomes and, related to the latter, that most of our patients had relatively low impairment levels and thus a very limited room for functional improvement. In any event, these results contrast with pre-clinical evidence demonstrating that BDNF is crucial in mediating the positive effects that CE has on functional stroke recovery.^28^

One possible explanation for the discrepancies between animal and human studies could be the different sources from which BDNF is typically measured across species. In animal models, BDNF can be measured directly in the brain, whereas in humans, it is measured peripherally, assuming its concentration reflects central neural processes. Previous studies suggest that BDNF can be transported unidirectionally from peripheral circulation to the brain by crossing the blood-brain barrier (BBB)^47^ and that the brain might be the primary source of circulating BDNF both at rest and during CE.^48^ This is supported by studies showing correlations between peripheral BDNF levels and central brain concentrations.^49^ However, this notion has been challenged by evidence indicating that neurotrophins, including BDNF, do not cross the BBB in large amounts unless they are conjugated with a molecular “Trojan horse”.^50^ When conjugated to a chimeric peptide, intravenous administration of BDNF reduces stroke volume and improves functional outcomes in rats with middle cerebral artery occlusion.^51^

This disparity between BDNF sources has also been observed during early post-stroke stages in animal models, where BDNF concentrations increase in the brain, while no changes are reported peripherally.^52^ This discrepancy underscores the need for caution in interpreting human studies and highlights the necessity for further studies to elucidate the role of circulating BDNF in central neural processes and its association with stroke recovery. Employing techniques such as positron emission tomography could provide more sensitive measures of brain BDNF utilization through the TrkB/BDNF system.^53^

Understanding the role of genetic variants on neuroplasticity biomarkers could help identify patients who are more likely to benefit from rehabilitation.^9^ It has been hypothesized that individuals with one or two copies of the met allele of the Val66Met polymorphism may show a decreased response to neuroplasticity-based interventions such as CE, due to diminished BDNF secretion.^54^ This study is the first to investigate the impact of the Val66Met polymorphism on serum BDNF levels in response to CE in individuals with stroke. In contrast to animal studies that consistently show the Val66Met polymorphism altering intracellular trafficking and activity-dependent BDNF expression, including in response to CE,^55^ BDNF_chronic_ and BDNF_acute_ were not influenced by Val66Met. Our findings align with other studies in neurotypical individuals reporting inconclusive results on the association between this genetic variant and BDNF levels following CE interventions,^24, 56, 57^ as well as its impact on post-stroke recovery outcomes.^58, 59^ Nevertheless, these findings should be interpreted with caution due to the large sample sizes typically required to detect true effects in genetic studies and the inherent variability of clinical stroke research.^60^

## CONCLUSION

This study is the largest trial investigating the effects of CE training on BDNF levels in individuals recovering from a stroke and the first trial exploring the effects in early subacute stages. Given that animal evidence suggests a period of heightened neuroplasticity and responsiveness to training during early stages of recovery,^1^ we expected a significant effect of CE on enhancing BDNF levels in early subacute stroke patients. Unexpectedly, our findings showed that despite significant improvements in cardiorespiratory fitness, CE had limited effects on both BDNF_chronic_ and BDNF_acute_. Similarly, BDNF responses were not associated with changes in cardiorespiratory fitness, recovery outcomes or influenced by BDNF Val66Met polymorphism. This aligns with previous studies that have not been able to establish a clear link between the upregulation of circulating BDNF following CE and stroke recovery improvements.^17^ Factors such as inflammation and stress responses during the early stages of post-stroke recovery, along with methodological and biological factors that contribute to increased variability, may have influenced these findings.

## AUTHOR CONTRIBUTIONS

Bernat de las Heras: design, data collection, data analysis, data interpretation, revisions, first draft. Lynden Rodrigues: data collection, data interpretation, revisions. Jacopo Cristini: data collection, data interpretation, revisions. Eric Yu: data analysis, data interpretation, revisions. Ziv Gan-Or: data analysis, data interpretation, revisions. Nathalie Arbour: data analysis, data interpretation, revisions. Alexander Thiel: data interpretation, revisions. Ada Tang: data interpretation, revisions. Joyce Fung: data interpretation, revisions. Janice J Eng: data interpretation, revisions. Marc Roig: conception, design, data collection, data analysis, data interpretation, revisions, supervision. All authors approved the final version of the manuscript and agree to be accountable for all aspects of the work in ensuring that questions related to the accuracy or integrity of any part of the work are appropriately investigated and resolved.

## DATA AVAILABILITY STATEMENT

Data is available upon reasonable request.

## REFERENCES

1. Murphy TH, Corbett D. Plasticity during stroke recovery: from synapse to behaviour. Nat Rev Neurosci. 2009;10(12):861–872. 10.1038/nrn2735

2. Carmichael ST. Cellular and molecular mechanisms of neural repair after stroke: Making waves. Ann Neurol. 2006;59(5):735–742. doi:10.1002/ana.20845

3. Bernhardt J, Hayward KS, Kwakkel G, et al. Agreed definitions and a shared vision for new standards in stroke recovery research: The Stroke Recovery and Rehabilitation Roundtable taskforce. Int J Stroke. Jul 2017;12(5):444–450. doi:10.1177/1747493017711816

4. Dromerick AW, Geed S, Barth J, et al. Critical Period After Stroke Study (CPASS): A phase II clinical trial testing an optimal time for motor recovery after stroke in humans. Proc Natl Acad Sci U S A. Sep 28 2021;118(39)doi:10.1073/pnas.2026676118

5. Lu B, Pang PT, Woo NH. The yin and yang of neurotrophin action. Nature Reviews Neuroscience. 2005;6(8):603–614. doi:10.1038/nrn1726

6. Clarkson AN, Overman JJ, Zhong S, Mueller R, Lynch G, Carmichael ST. AMPA Receptor-Induced Local Brain-Derived Neurotrophic Factor Signaling Mediates Motor Recovery after Stroke. The Journal of Neuroscience. 2011;31(10):3766–3775. doi:10.1523/jneurosci.5780-10.2011

7. MüLler HD, Hanumanthiah KM, Diederich K, Schwab S, SchäBitz W-RD, Sommer C. Brain-Derived Neurotrophic Factor But Not Forced Arm Use Improves Long-Term Outcome After Photothrombotic Stroke and Transiently Upregulates Binding Densities of Excitatory Glutamate Receptors in the Rat Brain. Stroke. 2008;39(3):1012–1021. doi:10.1161/strokeaha.107.495069

8. Pearson-Fuhrhop KM, Kleim JA, Cramer SC. Brain Plasticity and Genetic Factors. 2009;16(4):282–299. doi:10.1310/tsr1604-282

9. Stewart JC, Cramer SC. Genetic Variation and Neuroplasticity: Role in Rehabilitation After Stroke. J Neurol Phys Ther. Jul 2017;41 Suppl 3(Suppl 3 IV STEP Spec Iss):S17–s23. doi:10.1097/npt.0000000000000180

10. Mowla SJ, Pareek S, Farhadi HF, et al. Differential Sorting of Nerve Growth Factor and Brain-Derived Neurotrophic Factor in Hippocampal Neurons. The Journal of Neuroscience. 1999;19(6):2069–2080. doi:10.1523/jneurosci.19-06-02069.1999

11. Cotman C. Exercise: a behavioral intervention to enhance brain health and plasticity. Trends Neurosci. 2002;25(6):295–301. doi:10.1016/s0166-2236(02)02143-4

12. Ploughman M, Austin MW, Glynn L, Corbett D. The effects of poststroke aerobic exercise on neuroplasticity: a systematic review of animal and clinical studies. Transl Stroke Res. Feb 2015;6(1):13–28. doi:10.1007/s12975-014-0357-7

13. Knaepen K. Neuroplasticity – Exercise-Induced Response of Peripheral Brain-Derived Neurotrophic Factor. A Systematic Review of Experimental Studies in Human Subjects. Sports Med 2010;40(9):765–801. doi:10.2165/11534530-000000000-00000

14. Berchtold NC, Chinn G, Chou M, Kesslak JP, Cotman CW. Exercise primes a molecular memory for brain-derived neurotrophic factor protein induction in the rat hippocampus. Neuroscience. 2005;133(3):853–61. doi:10.1016/j.neuroscience.2005.03.026

15. Stanne TM, Åberg ND, Nilsson S, et al. Low Circulating Acute Brain-Derived Neurotrophic Factor Levels Are Associated With Poor Long-Term Functional Outcome After Ischemic Stroke. Stroke. 2016;47(7):1943–1945. doi:10.1161/strokeaha.115.012383

16. Ashcroft SK, Ironside DD, Johnson L, Kuys SS, Thompson-Butel AG. Effect of Exercise on Brain-Derived Neurotrophic Factor in Stroke Survivors: A Systematic Review and Meta-Analysis. Stroke. 2022;53(12):3706–3716. doi:10.1161/strokeaha.122.039919

17. De Las Heras B, Rodrigues L, Cristini J, et al. Measuring Neuroplasticity in Response to Cardiovascular Exercise in People With Stroke: A Critical Perspective. Neurorehabil Neural Repair. Jan 31 2024:15459683231223513. doi:10.1177/15459683231223513

18. Petryshen TL, Sabeti PC, Aldinger KA, et al. Population genetic study of the brain-derived neurotrophic factor (BDNF) gene. 2010;15(8):810–815. doi:10.1038/mp.2009.24

19. Mackay-Lyons M, Billinger SA, Eng JJ, et al. Aerobic Exercise Recommendations to Optimize Best Practices in Care After Stroke: AEROBICS 2019 Update. Phys Ther. 2019;doi:10.1093/ptj/pzz153

20. Billinger SA, Tseng BY, Kluding PM. Modified Total-Body Recumbent Stepper Exercise Test for Assessing Peak Oxygen Consumption in People With Chronic Stroke. 2008;88(10):1188–1195. doi:10.2522/ptj.20080072

21. Fletcher GF, Ades PA, Kligfield P, et al. Exercise Standards for Testing and Training. Circulation. 2013;128(8):873–934. doi:10.1161/cir.0b013e31829b5b44

22. Skriver K, Roig M, Lundbye-Jensen J, et al. Acute exercise improves motor memory: exploring potential biomarkers. Neurobiol Learn Mem. Dec 2014;116:46–58. doi:10.1016/j.nlm.2014.08.004

23. Polacchini A, Metelli G, Francavilla R, et al. A method for reproducible measurements of serum BDNF: comparison of the performance of six commercial assays. Sci Rep. Dec 10 2015;5:17989. doi:10.1038/srep17989

24. Lemos JR, Alves CR, De Souza SBC, et al. Peripheral vascular reactivity and serum BDNF responses to aerobic training are impaired by the BDNF Val66Met polymorphism. Physiol Genomics. 2016;48(2):116–123. doi:10.1152/physiolgenomics.00086.2015

25. Boyne P, Dunning K, Carl D, Gerson M, Khoury J, Kissela B. Within-session responses to high-intensity interval training in chronic stroke. Med Sci Sports Exerc. Mar 2015;47(3):476–84. doi:10.1249/MSS.0000000000000427

26. Nicolini C, Nelson AJ. Current Methodological Pitfalls and Caveats in the Assessment of Exercise-Induced Changes in Peripheral Brain-Derived Neurotrophic Factor: How Result Reproducibility Can Be Improved. Frontiers in Neuroergonomics. 2021;2doi:10.3389/fnrgo.2021.678541

27. De Havenon A, Sheth KN, Johnston KC, et al. Effect of Adjusting for Baseline Stroke Severity in the National Inpatient Sample. Stroke. 2021;52(11)doi:10.1161/strokeaha.121.035112

28. Ploughman M, Windle V, Maclellan CL, White N, Dore JJ, Corbett D. Brain-Derived Neurotrophic Factor Contributes to Recovery of Skilled Reaching After Focal Ischemia in Rats. Stroke. 2009;40(4):1490–1495. doi:10.1161/strokeaha.108.531806

29. Pandey A, Patel MR, Willis B, et al. Association Between Midlife Cardiorespiratory Fitness and Risk of Stroke. Stroke. 2016;47(7):1720–1726. doi:10.1161/strokeaha.115.011532

30. Luo L, Meng H, Wang Z, et al. Effect of high-intensity exercise on cardiorespiratory fitness in stroke survivors: A systematic review and meta-analysis. Ann Phys Rehabil Med. Jan 2020;63(1):59–68. doi:10.1016/j.rehab.2019.07.006

31. Ploughman M, Eskes GA, Kelly LP, et al. Synergistic Benefits of Combined Aerobic and Cognitive Training on Fluid Intelligence and the Role of IGF-1 in Chronic Stroke. Neurorehabilitation and Neural Repair. 2019;33(3):199–212. doi:10.1177/1545968319832605

32. Hsu CC, Fu TC, Huang SC, Chen CP-C, Wang J-S. Increased serum brain-derived neurotrophic factor with high-intensity interval training in stroke patients: A randomized controlled trial. Ann Phys Rehabil Med. 2021;64(4):101385. doi:10.1016/j.rehab.2020.03.010

33. Thacker JS, Xu Y, Tang C, Tupling AR, Staines WR, Mielke JG. A Single Session of Aerobic Exercise Mediates Plasticity-Related Phosphorylation in both the Rat Motor Cortex and Hippocampus. Neuroscience. Aug 1 2019;412:160–174. doi:10.1016/j.neuroscience.2019.05.051

34. Clarkson AN, Parker K, Nilsson M, Walker FR, Gowing EK. Combined Ampakine and BDNF Treatments Enhance Poststroke Functional Recovery in Aged Mice via AKT-CREB Signaling. Journal of Cerebral Blood Flow & Metabolism. 2015;35(8):1272–1279. doi:10.1038/jcbfm.2015.33

35. Boyne P, Meyrose C, Westover J, et al. Exercise intensity affects acute neurotrophic and neurophysiological responses poststroke. J Appl Physiol. 2019;126(2):431–443. doi:10.1152/japplphysiol.00594.2018

36. Szuhany KL, Bugatti M, Otto MW. A meta-analytic review of the effects of exercise on brain-derived neurotrophic factor. J Psychiatr Res. 2015;60:56–64. doi:10.1016/j.jpsychires.2014.10.003

37. Zalewska K, Ong LK, Johnson SJ, Nilsson M, Walker FR. Oral administration of corticosterone at stress-like levels drives microglial but not vascular disturbances post-stroke. Neuroscience. Jun 3 2017;352:30–38. doi:10.1016/j.neuroscience.2017.03.005

38. Kriz J, Lalancette-Hébert M. Inflammation, plasticity and real-time imaging after cerebral ischemia. Acta Neuropathol. 2009;117(5):497–509. doi:10.1007/s00401-009-0496-1

39. Schaaf MJ, De Kloet ER, Vreugdenhil E. Corticosterone effects on BDNF expression in the hippocampus. Implications for memory formation. Stress. May 2000;3(3):201–8. doi:10.3109/10253890009001124

40. Golia MT, Poggini S, Alboni S, et al. Interplay between inflammation and neural plasticity: Both immune activation and suppression impair LTP and BDNF expression. Brain Behav Immun. Oct 2019;81:484–494. doi:10.1016/j.bbi.2019.07.003

41. Chow LS, Gerszten RE, Taylor JM, et al. Exerkines in health, resilience and disease. Nature Reviews Endocrinology. 2022;18(5):273–289. doi:10.1038/s41574-022-00641-2

42. Cerqueira É, Marinho DA, Neiva HP, Lourenço O. Inflammatory Effects of High and Moderate Intensity Exercise—A Systematic Review. Front Physiol. 2020;10doi:10.3389/fphys.2019.01550

43. Carmichael ST, Archibeque I, Luke L, Nolan T, Momiy J, Li S. Growth-associated gene expression after stroke: evidence for a growth-promoting region in peri-infarct cortex. Exp Neurol. Jun 2005;193(2):291–311. doi:10.1016/j.expneurol.2005.01.004

44. Ploughman M, Granter-Button S, Chernenko G, et al. Exercise intensity influences the temporal profile of growth factors involved in neuronal plasticity following focal ischemia. Brain Res. 2007;1150:207–216. doi:10.1016/j.brainres.2007.02.065

45. Stoll P, Plessow A, Bratke K, Virchow JC, Lommatzsch M. Differential effect of clopidogrel and aspirin on the release of BDNF from platelets. J Neuroimmunol. Sep 15 2011;238(1-2):104–6. doi:10.1016/j.jneuroim.2011.06.015

46. Luo W, Liu T, Li S, et al. The Serum BDNF Level Offers Minimum Predictive Value for Motor Function Recovery After Stroke. Translational Stroke Research. Aug 2019;10(4):342–351. doi:10.1007/s12975-018-0648-5

47. Pan W. Transport of brain-derived neurotrophic factor across the blood–brain barrier Neuropharmacology 1998;37:1553–1561.

48. Rasmussen P, Brassard P, Adser H, et al. Evidence for a release of brain-derived neurotrophic factor from the brain during exercise. Exp Physiol. Oct 2009;94(10):1062–9. doi:10.1113/expphysiol.2009.048512

49. Klein AB, Williamson R, Santini MA, et al. Blood BDNF concentrations reflect brain-tissue BDNF levels across species. The International Journal of Neuropsychopharmacology. 2011;14(03):347–353. doi:10.1017/s1461145710000738

50. Pardridge WM. Blood-brain barrier drug targeting: the future of brain drug development. Mol Interv. Mar 2003;3(2):90–105, 51. doi:10.1124/mi.3.2.90

51. Zhang Y, Pardridge WM. Blood-brain barrier targeting of BDNF improves motor function in rats with middle cerebral artery occlusion. Brain Res. Sep 21 2006;1111(1):227–9. doi:10.1016/j.brainres.2006.07.005

52. Béjot Y, Mossiat C, Giroud M, Prigent-Tessier A, Marie C. Circulating and Brain BDNF Levels in Stroke Rats. Relevance to Clinical Studies. PLoS One. 2011;6(12):e29405. doi:10.1371/journal.pone.0029405

53. Bernard-Gauthier V, Boudjemeline M, Rosa-Neto P, Thiel A, Schirrmacher R. Towards tropomyosin-related kinase B (TrkB) receptor ligands for brain imaging with PET: radiosynthesis and evaluation of 2-(4-[(18)F]fluorophenyl)-7,8-dihydroxy-4H-chromen-4-one and 2-(4-([N-methyl-(11)C]-dimethylamino)phenyl)-7,8-dihydroxy-4H-chromen-4-one. Bioorg Med Chem. Dec 15 2013;21(24):7816–29. doi:10.1016/j.bmc.2013.10.012

54. Mang CS, Campbell KL, Ross CJ, Boyd LA. Promoting neuroplasticity for motor rehabilitation after stroke: considering the effects of aerobic exercise and genetic variation on brain-derived neurotrophic factor. Phys Ther. Dec 2013;93(12):1707–16. doi:10.2522/ptj.20130053

55. Ieraci A, Madaio AI, Mallei A, Lee FS, Popoli M. Brain-Derived Neurotrophic Factor Val66Met Human Polymorphism Impairs the Beneficial Exercise-Induced Neurobiological Changes in Mice. Neuropsychopharmacology. 2016;41(13):3070–3079. doi:10.1038/npp.2016.120

56. Helm EE, Matt KS, Kirschner KF, Pohlig RT, Kohl D, Reisman DS. The influence of high intensity exercise and the Val66Met polymorphism on circulating BDNF and locomotor learning. Neurobiol Learn Mem. Oct 2017;144:77–85. doi:10.1016/j.nlm.2017.06.003

57. Baird JF, Gaughan ME, Saffer HM, et al. The effect of energy-matched exercise intensity on brain-derived neurotrophic factor and motor learning. Neurobiology of Learning and Memory. 2018;156:33–44. doi:10.1016/j.nlm.2018.10.008

58. Cramer SC, See J, Liu B, et al. Genetic Factors, Brain Atrophy, and Response to Rehabilitation Therapy After Stroke. Neurorehabilitation and Neural Repair. 2021:154596832110628. doi:10.1177/15459683211062899

59. Shiner CT, Pierce KD, Thompson-Butel AG, Trinh T, Schofield PR, McNulty PA. BDNF Genotype Interacts with Motor Function to Influence Rehabilitation Responsiveness Poststroke. Front Neurol. 2016;7:69. doi:10.3389/fneur.2016.00069

60. Lee J-M, Fernandez-Cadenas I, Lindgren AG. Using Human Genetics to Understand Mechanisms in Ischemic Stroke Outcome: From Early Brain Injury to Long-Term Recovery. Stroke. 2021;52(9):3013–3024. doi:10.1161/strokeaha.121.032622

