## Supplementary material for "Investigating the Effect of Cardiovascular Exercise on Brain-Derived Neurotrophic Factor in Early Subacute Stroke": Suplementary Materials

**Supplementary Materials**

|  | MICT | HIIT | Total |
| --- | --- | --- | --- |
| HR_max_ (%) | 82.35 ± 8.06 | 81.89 ± 6.94 | 82.13 ± 7.05 |
| PPO _­_(%) | 63.69 ± 8.74 | 67.49 ± 13.74 | 65.59 ± 10.96 |
| Total Steps | 29384 ± 5932 | 20237 ± 4366 | 49464 ± 10183 |
| RPE (0-10) | 4.58 ± 1.31 | 5.15 ± 1.61 | 4.86 ± 1.35 |

**Supplementary Table 1**. Internal and External Training Load for the CE+standard care group. CE+standard care group's average internal and external training loads during both MICT and HIIT periods, including the warm-up and cool-down phases of each session. The average percentages of HR_max_ and PPO achieved during both MICT and HIIT periods were calculated based on VO_2_peak values at T0 and T1, respectively. Values are presented as mean and SD. Abbreviations: CE, cardiovascular exercise; HIIT, High-intensity interval training; HR, heart rate; MICT, Moderate-to-vigorous Continuous Training; PPO, peak power output; RPE, rate of perceived exertion.

|  | DFNum | DFDen | F ratio | p value |
| --- | --- | --- | --- | --- |
| BDNF_acute_T0 |  |  |  |  |
| Age | 1 | 61.5 | 0.02 | 0.876 |
| Sex | 1 | 61.5 | 0.23 | 0.630 |
| NIHSS | 1 | 61.5 | 0.19 | 0.664 |
| BMI | 1 | 61.6 | 3.02 | 0.087 |
| Val66Met | 1 | 61.6 | 0.009 | 0.923 |
| Time[Val66Met] | 6 | 254.0 | 1.48 | 0.185 |
| BDNF_chronic_ |  |  |  |  |
| Age | 1 | 62.0 | 0.08 | 0.775 |
| Sex | 1 | 62.5 | 0.76 | 0.386 |
| NIHSS | 1 | 62.8 | 0.30 | 0.585 |
| BMI | 1 | 65.5 | 2.10 | 0.152 |
| Val66Met | 1 | 62.3 | 0.07 | 0.793 |
| Group | 1 | 62.3 | 0.44 | 0.507 |
| Time | 2 | 172.0 | 1.87 | 0.157 |
| Time*Group[Val66Met] | 4 | 172.0 | 0.11 | 0.978 |
| BDNF_acute_ |  |  |  |  |
| Age | 1 | 60.1 | 0.18 | 0.673 |
| Sex | 1 | 62.4 | 3.15 | 0.081 |
| NIHSS | 1 | 62.4 | 0.004 | 0.952 |
| BMI | 1 | 71.9 | 0.11 | 0.737 |
| Val66Met | 1 | 61.2 | 0.61 | 0.436 |
| Group | 1 | 60.9 | 1.92 | 0.170 |
| Time | 2 | 171.0 | 2.67 | 0.072 |
| Time*Group[Val66Met] | 4 | 171.0 | 0.75 | 0.554 |

**Supplementary Table 2** Adjusted linear mixed models examining the influence of Val66Met polymorphism on acute BDNF changes at T0, as well as chronic and acute changes following cardiovascular exercise training. Abbreviations: BMI, body mass index; NIHSS, national institutes of health stroke scale. * p<0.05

|  | CE+standard care | | | Standard care | | |
| --- | --- | --- | --- | --- | --- | --- |
|  | **Estimate (95% CI)** | **p value** | **R^2^** | **Estimate (95% CI)** | **p value** | **R^2^** |
| UL-FMA T0-T2 |  |  |  |  |  |  |
| BDNF_chronic_  BDNF_acute_ | -7.45 (-0.0004, 0.0002)  0.0001 (-0.0002, 0.0006) | 0.661  0.411 | 0.20  0.22 | -6.06 (-0.0004, 0.0002)  0.0001 (-0.0003, 0.0006) | 0.717  0.464 | 0.38  0.38 |
| BBT T0-T2 |  |  |  |  |  |  |
| BDNF_chronic_  BDNF_acute_ | -0.0005 (-0.0001, -5.81)  -0.0001 (-0.0004, 0.0008) | 0.209  0.588 | 0.16  0.04 | -0.0004 (-0.0009, 9.79)  0.0003 (-0.0004, 0.001) | 0.103  0.325 | 0.62  0.55 |
| CRF T0-T2 |  |  |  |  |  |  |
| BDNF_chronic_  BDNF_acute_ | -2.91 (-0.0002, 0.0001)  -0.0001 (-0.0001, 0.0003) | 0.770  0.376 | 0.19  0.20 | -0.0001 (-0.0001, 0.0003)  6.55 (-0.0003, 0.0004) | 0.424  0.742 | 0.17  0.14 |

**Supplementary Table 3.** Adjusted multivariate linear regression examining associations between chronic and acute BDNF changes and changes in recovery outcomes, including upper-limb impairment, function, and cardiorespiratory fitness. Abbreviations: BBT, Box and Blocks Test; CE, cardiovascular exercise; CRF, cardiovascular fitness; UL-FMA, upper-limb Fugl-Meyer assessment. * p<0.05
